# Rifampicin monoresistant tuberculosis: exploring the interplay with HIV and mortality: an individual participant data meta-analysis

**DOI:** 10.1101/2025.03.26.25324718

**Authors:** Anna L. Jazza, Eskedar Getie Mekonnen, Helen Cox, Annelies Van Rie

**Affiliations:** Department of Family Medicine and Population Health, Global Health Institute, Faculty of Medicine and Health Sciences, University of Antwerp, Antwerp, Belgium; Adelaide Medical School, University of Adelaide, South Australia; Burnet Institute, Melbourne, Australia; Division of Medical Microbiology and Institute of Infectious Disease and Molecular Medicine, University of Cape Town, South Africa

## Abstract

Multidrug-resistant or rifampicin-resistant tuberculosis (MDR/RR-TB) poses a significant global public health challenge. While most people with RR-TB have MDR-TB (resistance to isoniazid and rifampicin), rifampicin mono-resistant tuberculosis (RmR-TB, rifampicin resistance, isoniazid susceptibility) is prevalent in certain regions. We aimed to compare mortality between patients with RmR-TB and MDR-TB and assess potential effect modification by HIV status during treatment.

We conducted an individual patient data meta-analysis involving 16,651 individuals with confirmed drug-resistant tuberculosis, all of whom had drug susceptibility test results for at least rifampicin, isoniazid, and fluoroquinolones and had initiated TB treatment. Mixed-effects logistic regression was used to estimate mortality during two-year treatment, adjusting for age, sex, history of TB, disease site, and HIV status. Stratified analyses explored effect modification by HIV status.

Among 16,568 patients, 2878 (17.4%) were classified as having RmR-TB, 11,236 (67.8%) as MDR-TB, 2384 (14.4%) as pre-XDR-TB, and 70 (0.4%) as XDR-TB. During the first four months of treatment, the cumulative incidence of mortality was higher among patients with RmR-TB compared to those with MDR-TB, pre-XDR-TB and XDR-TB. Patients with RmR-TB had 14% higher odds of mortality compared to patients with MDR-TB (32.2% vs 23.7%; aOR 1.14; 95% CI: 1.02–1.27), but a lower mortality compared to patients with pre-XDR or XDR-TB. Although HIV status did not significantly modify the effect (p-value interaction = 0.362), the odds of mortality was only higher among RmR-TB patients with HIV co-infection (aOR 1.17; 95% CI:1.03–1.33).

Using a large individual patient data meta-analysis, this study confirmed the counterintuitive finding of prior smaller studies that the odds of mortality are higher among patients with RmR-TB compared to MDR-TB. While HIV co-infection was not an effect modifier, the higher odds of mortality were only observed among RmR TB patients living with HIV.

## Introduction

Drug-resistant tuberculosis (DR-TB) is a significant public health concern, impeding progress towards TB control. Multidrug resistant or rifampicin-resistant TB (MDR/RR-TB) accounts for approximately 9.4% of the global TB burden, with an estimated 400,000 persons with new TB infection in 2023[1]. While most people with MDR/RR-TB have MDR-TB, defined as resistance to both rifampicin (RIF) and isoniazid (INH), many regions report a growing burden of rifampicin mono-resistant TB (RmR-TB), defined as resistance to RIF but susceptible to INH [2–6].

The advent of molecular diagnostic tools such as Xpert MTB/RIF has facilitated the rapid routine diagnosis of RR-TB [3, 7–9]. Additionally, access to the Hain GenoType MTBDR line probe assay, the Xpert MTB/XDR cartridge and a growing number of other cartridge-based assays now facilitated the differentiation of RR-TB as RmR-TB or MDR-TB in routine clinical settings [10–12].

Studies have found that TB patients living with HIV are more likely to develop RmR-TB [3, 13, 14]. In South Africa, where 66% of TB patients are co-infected with HIV, an estimated 23.8% of these patients diagnosed with RR-TB have RmR-TB [3]. Although it may seem counterintuitive, several studies have indicated that RmR-TB poses a higher risk of mortality than MDR-TB. While HIV co-infection has been proposed as a potential effect modifier of this effect, the underlying reasons for the higher mortality among people with RmR-TB are not yet fully understood [2, 3, 6, 14–17].

To better quantify whether mortality with RmR-TB is higher than that of MDR-TB and test the hypothesis that HIV modifies the effect of RmR-TB on mortality, we performed a large individual participant data meta-analysis. A more comprehensive understanding of the mortality associated with RmR-TB could inform strategies for improving treatment outcomes among people with RmR-TB and help prevent the transmission of RmR *Mycobacterium tuberculosis* strains [4, 15, 18].

## Materials and methods

### Data sources, inclusion and exclusion criteria

A systematic review was conducted by searching articles in PubMed/MEDLINE, Web of Science, and the Cochrane Library. The search terms employed were:

(*Mycobacterium Tuberculosis* OR *mtb* OR tb OR tuberculosis) AND ((drug-resistant tuberculosis OR multi-drug-resistant tuberculosis OR MDR-TB or MDR/RR-TB OR (Rifampicin resistant Tuberculosis OR RR-TB OR rifampicin resistance OR rifampicin mono-resistance OR RmR-TB OR Pre-extensively resistant TB OR pre-XDRTB OR XDRTB OR extensively resistant TB)). The protocol for this review was registered in PROSPERO (registration number CRD42023487293).

Studies were eligible if they provided *Mtb* drug susceptibility testing (DST) results at treatment initiation. The initial search identified several eligible articles that had been included in a prior systematic review [19]. The data from studies in this prior review had been pooled into the TB treatment individual patient data platform (TB-IPD), a collaborative initiative led by the World Health Organization (WHO), Global TB Programme, and University College London (UCL) [20, 21]. Additionally, we identified the TB Portals Consortium, a platform designed for sharing de-identified DR-TB data collected by various global institutions [22].

The authors from three eligible studies identified [14, 23, 24] and the data access committees for the TB-IPD team and TB Portals Consortium agreed to share de-identified individual data following a formal data-sharing agreement. To avoid overlap between datasets, we requested data from the 2018 TB-IPD database.

Data was requested for the following variables: age, sex, body mass index (BMI), country of residence, HIV status, previous TB history, site of the disease, year of TB diagnosis, DST results, prescribed TB regimen, treatment duration, treatment outcome, date started treatment (or year), and the date the outcome was observed.

Patients were included in the individual patient data (IPD) meta-analysis if they had initiated TB treatment, had a treatment outcome recorded, and had DST results for at least INH, RIF and fluoroquinolones (FQ) at treatment initiation. Patients with MDR-TB were excluded if DST data for FQ were missing. In cases where DST results for bedaquiline, linezolid, or pretomanid were not available, strains were considered susceptible to those drugs, as most included studies preceded the use of these drugs in routine care. Patients whose treatment outcome was missing, categorized as ’transferred out’ or ’still on treatment’, were also excluded from the analysis.

### Data analysis

To create a unified analytic database, we first cleaned each individual dataset and assessed the datasets for missing data for potential confounding variables (age, sex history of TB, site of the disease (extra-pulmonary TB (EPTB), pulmonary TB (PTB), or both) and effect modifier (HIV status) as identified by a directed acyclic graph (DAG) (S1 Fig). We examined the pattern of missingness and evaluated the association between patients with observed values and those with missing observations. Little’s test was used to assess missing data mechanism for data missing completely at random.

The drug resistance profile of each patient was determined based on molecular and phenotypic DST results. Patients were classified as RmR if the *Mtb* strain was resistant to RIF and susceptible to INH; as MDR-TB if resistant to RIF and INH but susceptible to FQ; pre-extensively drug-resistant (pre-XDR) if resistant to RIF, INH, and FQ; and XDR if also resistant to bedaquiline (BDQ) and/or linezolid (LZD)[25]. The primary outcome mortality, was censored at 24 months follow-up given the maximum WHO recommended DR-TB treatment duration of 18–20 months [26]. Treatment regimens were classified as FQ, BDQ, and/or LZD containing regimens. We also classified treatment regimens by the number of WHO Group A drugs namely FQ (levofloxacin, moxifloxacin) BDQ, and LZD included[26].

All analyses were performed using R statistical software version 4.4.0. We derived descriptive statistics for the continuous and categorical variables using frequencies, mean and standard deviation (SD), or median and interquartile range (IQR). The difference in proportions was determined with a chi-squared test statistic. A one-stage IPD meta-analysis was conducted, clustering patients by country to account for potential variations in RR-TB management, healthcare infrastructure, and socio-economic factors that could influence treatment outcomes.

In the primary analysis, we performed a complete case analysis comparing mortality between patients with RmR-TB and those with MDR-TB. We used a mixed-effects logistic regression model to derive the crude and adjusted odds ratio (OR) and their 95% confidence interval (95% CI) for the association between mortality and type of RR-TB (RmR-TB versus MDR-TB). Patients with an outcome of loss to follow-up (LTFU) were excluded from this analysis.

To evaluate effect modification by HIV status, we assessed the joint effects of HIV status and RmR-TB by incorporating an interaction term in the mixed-effects logistic regression model. We tested the significance of the interaction effect using likelihood ratio tests (with a significance threshold of p < 0.05) and derived stratified effect estimates (adjusted odds ratios (aOR) along with their 95% CI) within each stratum of HIV status.

Several sensitivity analyses were performed to ensure the robustness of our analysis. To account for LTFU as a competing risk, we generated cumulative Incidence functions (CIFs) to estimate the two-year mortality rate and applied a sub-distribution hazard regression model to estimate the sub-distribution hazard ratios (sdHR) and their 95% CI. In addition to comparing RmR-TB to MDR-TB we also compared two-year mortality of RmR-TB vs MDR-TB, pre-XDR and XDR-TB, using mixed effect logistic regression as well as sub-distribution hazard regression models.

### Ethics statement

We obtained formal permission to use data from the data access committee of the databases and study investigators of the datasets included in this study.

## Results

### Study participants characteristics

The two databases (TB portals and TB-IPD) combined with the data from three published studies resulted in 40,022 eligible patients. Of these, 3.6% (n= 1455) were excluded due to missing DST results and 85 patients from the Cox cohort [14] whose year of diagnosis was between 2014 and 2015 to avoid overlap with the Schnippel cohort [23] (Fig 1).

**Fig 1:**
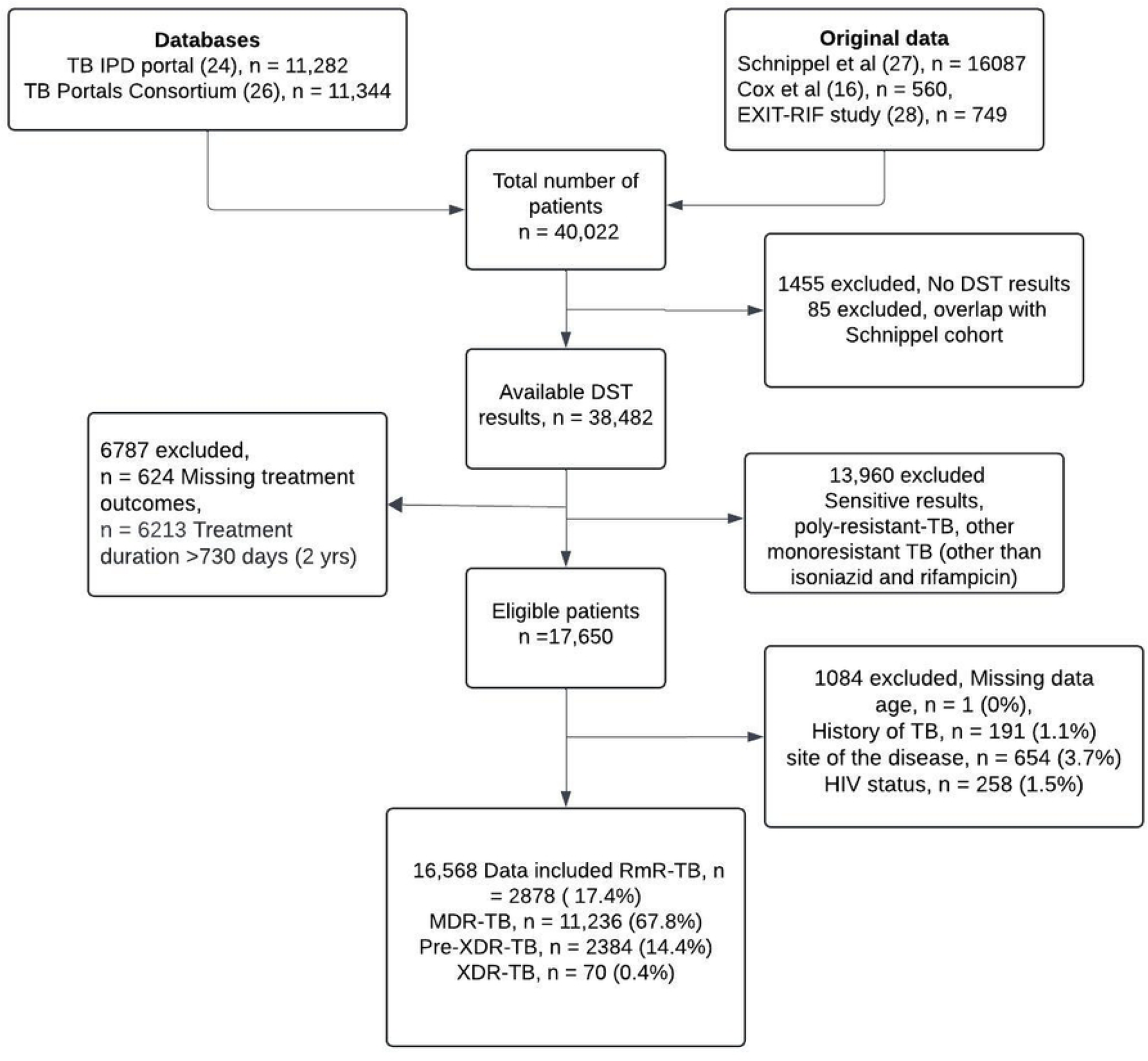
Flow chart of study participants and data sources included in this study DST = Drug susceptibility testing, TB-IPD= Tuberculosis treatment individual patient data, RmR-TB=Rifampicin monoresistant tuberculosis, MDR-TB = Multi-drug resistant tuberculosis, Pre-XDR-TB=pre-extensively resistant tuberculosis, XDR-TB=extensively resistant tuberculosis.

Of the remaining 38,482 patients with DST results, 13,960 patients were excluded as they were diagnosed with either drug-sensitive TB, poly-resistant TB, or mono-resistant TB other than RmR-TB. An additional 624 patients were excluded due to missing treatment outcome, and 6213 were excluded because they either did not start treatment or received treatment for more than two years.

Of the 16,568 patients included in the final analysis, 17.4% (2,878) were classified as having RmR-TB, 67.8% (11,236) as MDR-TB, 14.4% (2384) as pre-XDR-TB and 0.4% (70) as XDR-TB. Just over half (54.2%, n = 8,979) had a successful treatment outcome with 45.1% (7,466) cured and 9.1% (1,513) completing treatment. Over the course of treatment, 21% (3,483) patients died, 19.1% (3,169) were LTFU, and 5.7% (937) experienced treatment failure.

Of the 16,568 IPD participants, 2454 were again excluded from the primary analysis comparing RmR-RB with MDR-TB because they were classified as pre-XDR-TB or XDR-TB, and 2851 were further excluded because their treatment outcome was LTFU. The remaining 11,287 patients resided in four WHO regions, with the African region, primarily South Africa contributing the majority (68%, n = 7627) of the data.

The mean (SD) age of participants was 38.4 (12.4) years, 59% were male, 51% were living with HIV, 51% had a history of TB, and almost all (96%) had pulmonary TB. The median (IQR) duration of treatment was 18.4 (12) months, indicating that most patients received the long regimen.

Almost all (95%) patients received a FQ-containing regimen, 19.3% a BQD-containing regimen, and 10% received a LZD-containing regimen. Most (64%) participants were treated with only one group A drug, 8% received two group A drugs, 15% three group A drugs, and the rest 12% did not receive any group A drug (Table 1).

**Table 1.** Characteristics of RmR-RB and MDR-TB patients included in the primary analysis stratified by resistance type

| Variable | Resistance type |  | Overall (N = 11,287) |
| --- | --- | --- | --- |
|  | RmR-TB (N = 2163) | MDR-TB (N = 9124) |  |
| Age (years) |  |  |  |
| Mean (SD) | 36.9 (11.5) | 38.8 (12.6) | 38.4 (12.4) |
| Treatment duration (days) |  |  |  |
| Median (IQR) | 526 [517.5] | 566 [359] | 560 [365] |
| Sex |  |  |  |
| Female | 946 (43.7%) | 3678 (40.3%) | 4624 (41.0%) |
| Male | 1217 (56.3%) | 5446 (59.7%) | 6663 (59.0%) |
| Site of the disease |  |  |  |
| EPTB | 63 (2.9%) | 96 (1.1%) | 159 (1.4%) |
| PTB | 2088 (96.5%) | 8739 (95.8%) | 10827 (95.9%) |
| Both | 12 (0.6%) | 289 (3.2%) | 301 (2.7%) |
| History of TB |  |  |  |
| No | 1071 (49.4%) | 4509 (49.4%) | 5580 (49.4%) |
|  | RmR-TB (N = 2163) | MDR-TB (N = 9124) |  |
| Yes | 1092(50.5%) | 4615 (50.6%) | 5707 (50.6%) |
| Mortality |  |  |  |
| Alive | 1463 (67.6%) | 6958 (76.3%) | 8421 (74.6%) |
| Dead | 700 (32.4%) | 2166 (23.7%) | 2881 (25.4%) |
| HIV status |  |  |  |
| Negative | 505 (23.3%) | 4979 (54.6%) | 5484 (48.6%) |
| Positive | 1658 (76.7%) | 4145 (45.4%) | 5803 (51.4%) |
| WHO region |  |  |  |
| African | 2124 (98.2%) | 5503 (60.3%) | 7627 (67.6%) |
| Americas | 0 (0%) | 244 (2.7%) | 244 (2.2%) |
| European | 37 (1.7%) | 2550 (27.9%) | 2587 (22.9%) |
| Western Pacific | 2 (0.1%) | 827 (9.1%) | 829 (7.3%) |
| Bedaquiline containing regimen |  |  |  |
| No | 1981 (91.6%) | 7116 (78.0%) | 9097 (80.6%) |
| Yes | 182 (8.4%) | 2008 (22.0%) | 2190 (19.4%) |
| Fluoroquinolone containing regimen<sup>a</sup> |  |  |  |
| No | 44 (2.1%) | 484 (5.3%) | 528 (4.7%) |
| Yes | 2119 (98.0%) | 8640 (94.7%) | 10759 (95.3%) |
| Linezolid containing regimen |  |  |  |
| No | 2038 (94.2%) | 6989 (76.6%) | 9027 (80.0%) |
| Yes | 125 (5.8%) | 2135 (23.4%) | 2260 (10.0%) |
| Number of group A drugs<sup>b</sup> |  |  |  |
| 0 | 94 (4.3%) | 1245 (13.6%) | 1339 (11.9%) |
| 1 | 1874 (86.6%) | 5389 (59.1%) | 7263 (64.3%) |
| 2 | 64 (3.0%) | 898 (9.8%) | 962 (8.5%) |
| 3 or more | 131 (6.1%) | 1592 (17.4%) | 1723 (15.3%) |

<sup>a</sup>Fluoroquinolones include levofloxacin, moxifloxacin, ofloxacin, moxifloxacin, gatifloxacin
<sup>b</sup>WHO Group A drugs include levofloxacin, moxifloxacin, bedaquiline and linezolid
SD = Standard deviation, IQR = Interquartile range, EPTB = extra-pulmonary tuberculosis, PTB = pulmonary tuberculosis, RmR-TB = Rifampicin monoresistant tuberculosis, MDR-TB = Multi drug-resistant tuberculosis

### Mortality among patients with RmR-TB and MDR-TB during treatment

Higher mortality was observed among RmR-TB compared to MDR-TB patients (32.4% vs. 23.7%; p < 0.001). In multivariable analysis, increasing age, a history of TB infection, diagnosis of RmR-TB (vs MDR-TB), and HIV positivity were associated with mortality during treatment. Patients with RmR-TB had 14% higher odds of mortality than those with MDR-TB (aOR 1.14; 95% CI:1.02–1.27; p-value = 0.018). Patients with HIV/TB co-infection had nearly double the odds of mortality compared to their HIV negative counterparts (aOR 1.94; 95% CI: 1.73 – 2.17; p-value <0.001), and patients with a history of TB had 45% higher odds of mortality (aOR 1.45, 95% CI: 1.32, 1.59; p-value <0.001) than those not previously treated for TB. Among the previously treated patients with HIV co-infection, the odds of mortality were 25% higher among RMR-TB patients than those with MDR-TB (aOR 1.25; CI 1.05 - 1.41; p-value = 0.011). In contrast, no significant difference was observed among patients not previously treated for TB (aOR 1.04; CI 0.88 - 1.23; p-value = 0.6). Considering LTFU as a competing risk, the sub-distribution mortality hazard during treatment was 11% higher among patients with RmR-TB compared to those with MDR-TB (sdHR 1.11; 95 % CI 1.02–1.22). Among previously treated patients, the odds of mortality were 45% higher compared to those who had never been treated for TB (aOR 1.45; 95% CI 1.32–1.59; p < 0.001) (Table 2).

**Table 2:** Mortality (as a treatment outcome) among patients with rifampicin mono-resistant and multidrug-resistant tuberculosis.

| Variable | Multivariable logistic regression |  |  | Sub-distribution hazard model |  |
| --- | --- | --- | --- | --- | --- |
|  | aOR | 95% CI | p-value | sdHR | 95% CI |
| Age | 1.03 | 1.02 – 1.03 | <0.001 | 1.03 | 1.02 – 1.03 |
| Sex |  |  |  |  |  |
| Female | Ref |  |  | Ref |  |
| Male | 1.04 | 0.95 – 1.15 | 0.369 | 0.96 | 0.89 – 1.04 |
| History of TB |  |  |  |  |  |
| No | Ref |  |  | Ref |  |
| Yes | 1.45 | 1.32 – 1.59 | <0.001 | 1.24 | 1.15 – 1.33 |
| Site of the disease |  |  |  |  |  |
| EPTB | Ref |  |  | Ref |  |
| PTB | 0.61 | 0.45 – 0.82 | 0.065 | 0.93 | 0.69–1.25 |
| Both | 1.55 | 0.97 – 2.47 | 0.040 | 1.08 | 0.74 – 1.58 |
| Resistance type |  |  |  |  |  |
| MDR-TB | Ref |  |  | Ref |  |
| RmR-TB | 1.14 | 1.02 – 1.27 | 0.018 | 1.11 | 1.02 – 1.22 |
| HIV status |  |  |  |  |  |
| Negative | Ref |  |  | Ref |  |
| Positive | 1.94 | 1.73 – 2.17 | <0.001 | 2.30 | 2.12–2.49 |

Comparing results from a multivariable logistic regression and sub-distribution hazard model.
aOR = adjusted Odds Ratio, CI = Confidence Interval, sdHR=Sub-distribution Hazard Ratio,
EPTB = extra pulmonary tuberculosis, PTB = pulmonary tuberculosis, Both = extra pulmonary
tuberculosis and pulmonary tuberculosis, RmR-TB = Rifampicin monoresistant tuberculosis,

Comparing patient survival censoring LTFU, the cumulative incidence rate of mortality was higher for patients with RmR-TB compared to MDR-TB. Within the first 100 days of treatment, 13% of patients with RmR-TB had died, compared to 9% of those with MDR-TB. By one year, mortality increased to 21% for RmR-TB and 16% for MDR-TB. At two years, 30% of RmR-TB patients had died, compared to 25% of those with MDR-TB (Fig 2).

**Fig 2:**
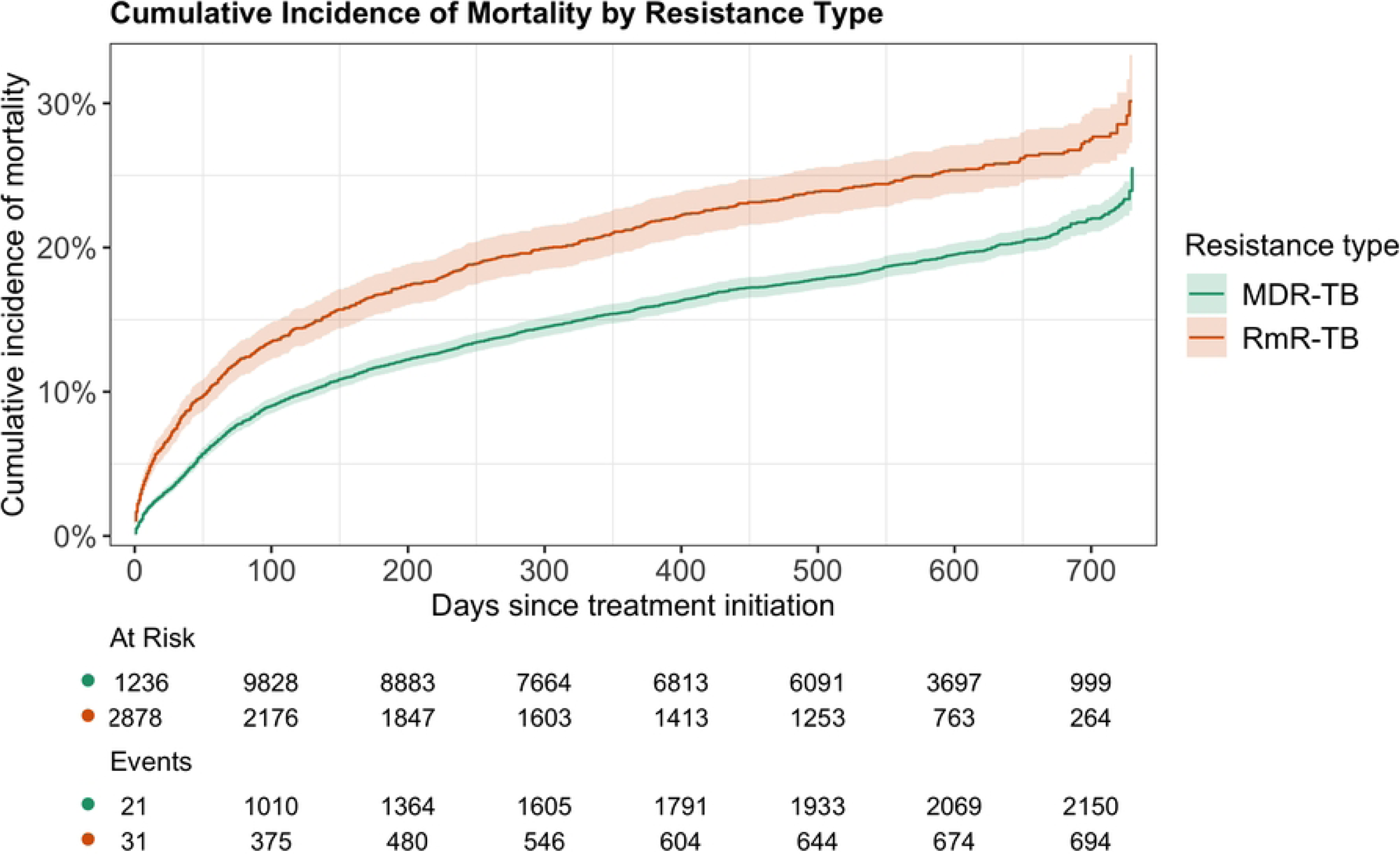
Cumulative incidence of two-year mortality stratified by resistance type. RmR-TB = Rifampicin monoresistant tuberculosis, MDR-TB = multi-drug-resistant tuberculosis

### Effect Modification by HIV

The proportion of HIV/TB co-infection was higher among RmR-TB patients compared to MDR-TB patients (76.9% vs. 45.5%; p < 0.001). The stratified analysis revealed a differential impact of RR-TB type on mortality. Among those with HIV/TB co-infection, patients with RmR-TB had 17% higher odds of mortality than those with MDR-TB (aOR 1.17; 95% CI:1.03–1.32; p-value = 0.013). In contrast, mortality among HIV-negative patients was similar for both RmR-TB and MDR-TB patients (aOR 0.99; 95% CI 0.77–1.27; p-value >0.9). However, the interaction effect between HIV and RmR-TB on mortality was not statistically significant (p-value= 0.387) (Fig 3).

**Fig 3:**
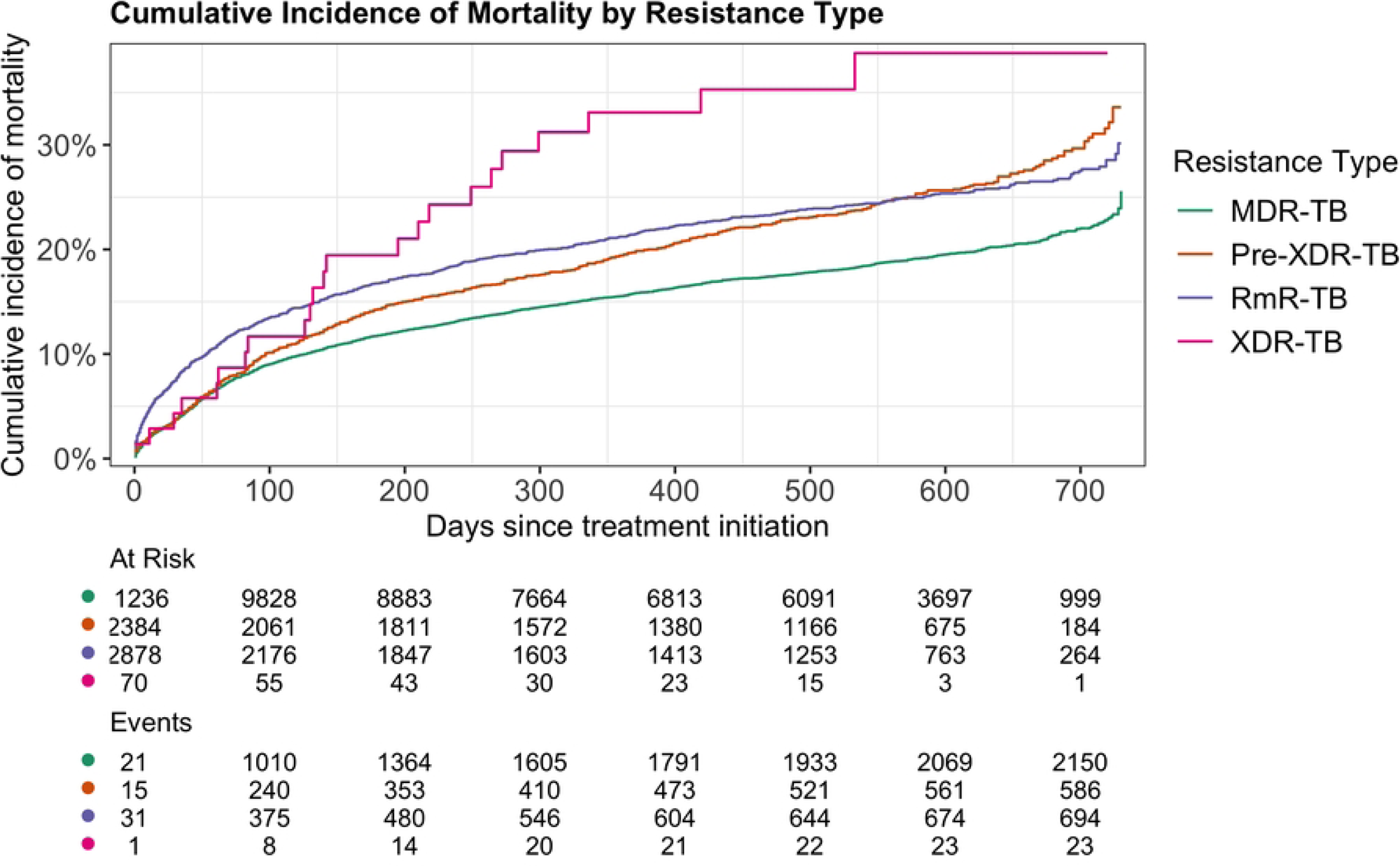
Effect of RmR-TB versus MDR-TB on mortality stratified by HIV status. aOR = Adjusted Odds Ratio, CI = Confidence Interval

### Mortality of RmR-TB compared to Pre-XDR-TB and XDR-TB

Patients with RmR-TB had 56% lower odds of two-year mortality compared to those with Pre-XDR-TB (aOR 0.44; 95% CI 0.36 – 0.52; p-value < 0.001). Accounting for LTFU as a competing risk, the sub-distribution hazard of mortality was 28% lower among patients with RmR-TB compared to those with Pre-XDR-TB (sdHR = 0.72; 95% CI 0.63 – 0.82). Similarly, patients with RmR-TB had 59% lower odds of two-year mortality compared to those with XDR-TB (aOR 0.41; 95% CI 0.18 – 0.93; p-value = 0.033). The sub-distribution hazard of mortality was 34% lower among patients with RmR-TB compared to those with XDR-TB (sdHR = 0.66; 95% CI 0.44 – 1.00) Table 3 and Table 4.

**Table 3:**
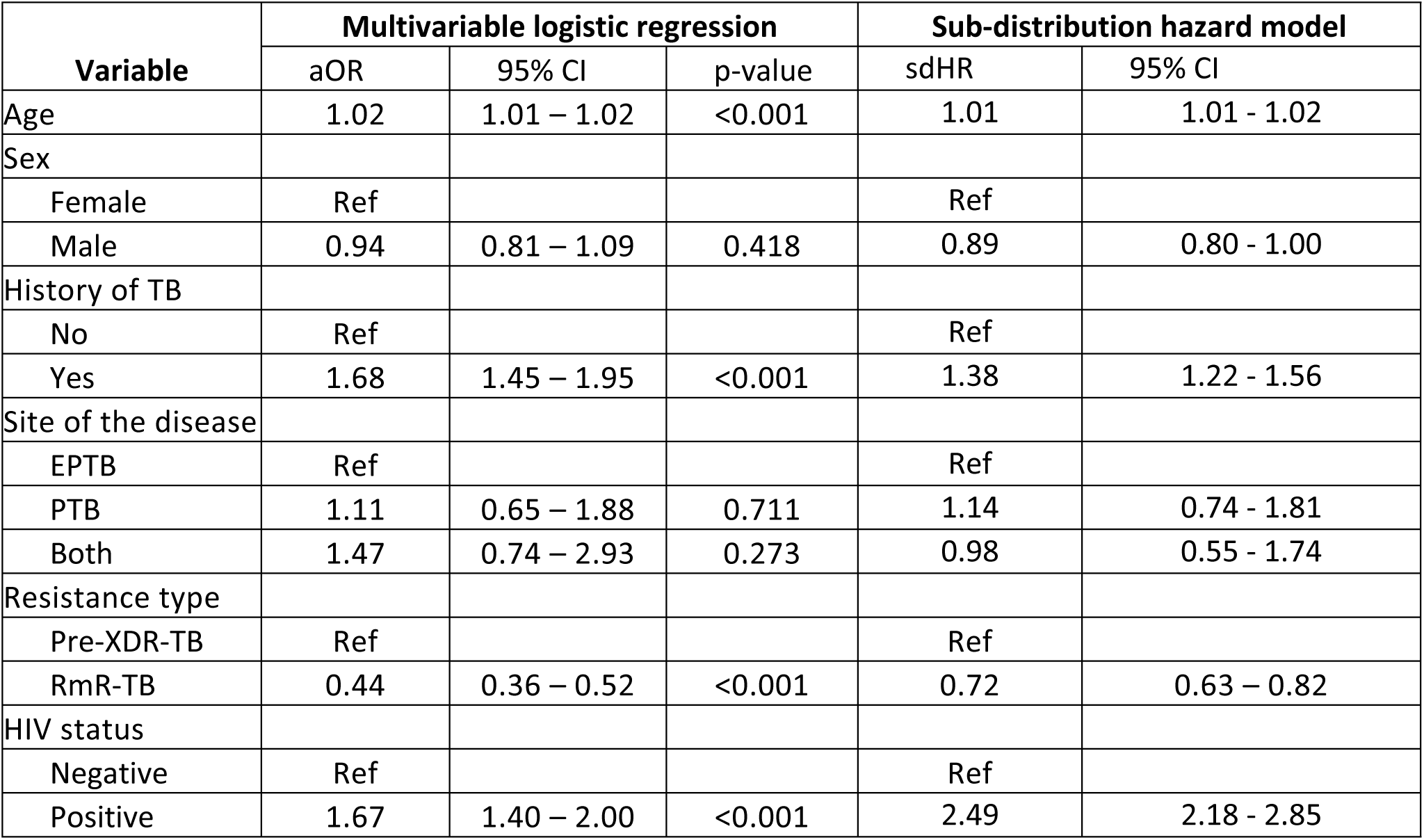
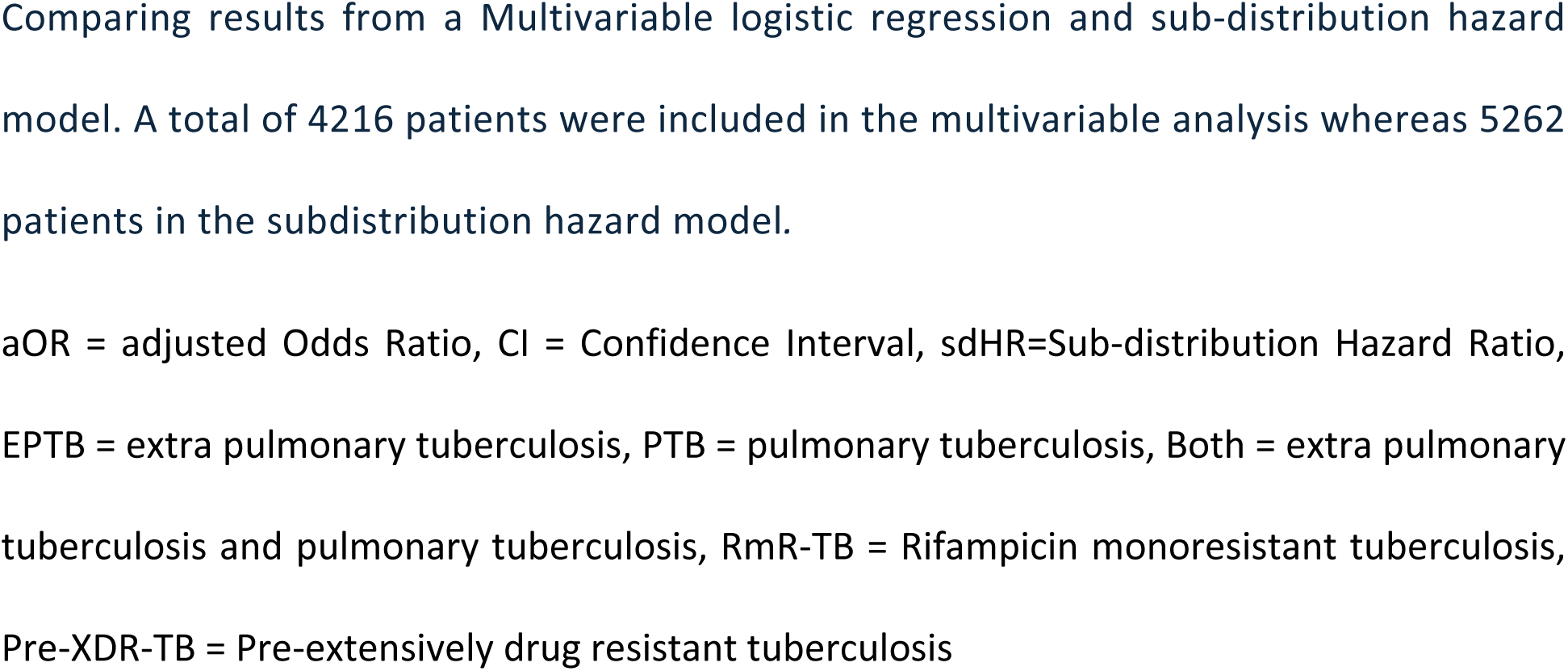
Two-year mortality among patients with rifampicin mono-resistant and pre-extensively drug-resistant tuberculosis.

| Variable | Multivariable logistic regression |  |  | Sub-distribution hazard model |  |
| --- | --- | --- | --- | --- | --- |
|  | aOR | 95% CI | p-value | sdHR | 95% CI |
| Age | 1.02 | 1.01 – 1.02 | <0.001 | 1.01 | 1.01 - 1.02 |
| Sex |  |  |  |  |  |
| Female | Ref |  |  | Ref |  |
| Male | 0.94 | 0.81 – 1.09 | 0.418 | 0.89 | 0.80 - 1.00 |
| History of TB |  |  |  |  |  |
| No | Ref |  |  | Ref |  |
| Yes | 1.68 | 1.45 – 1.95 | <0.001 | 1.38 | 1.22 - 1.56 |
| Site of the disease |  |  |  |  |  |
| EPTB | Ref |  |  | Ref |  |
| PTB | 1.11 | 0.65 – 1.88 | 0.711 | 1.14 | 0.74 - 1.81 |
| Both | 1.47 | 0.74 – 2.93 | 0.273 | 0.98 | 0.55 - 1.74 |
| Resistance type |  |  |  |  |  |
| Pre-XDR-TB | Ref |  |  | Ref |  |
| RmR-TB | 0.44 | 0.36 – 0.52 | <0.001 | 0.72 | 0.63 – 0.82 |
| HIV status |  |  |  |  |  |
| Negative | Ref |  |  | Ref |  |
| Positive | 1.67 | 1.40 – 2.00 | <0.001 | 2.49 | 2.18 - 2.85 |

Comparing results from a Multivariable logistic regression and sub-distribution hazard model. A total of 4216 patients were included in the multivariable analysis whereas 5262 patients in the subdistribution hazard model.
aOR = adjusted Odds Ratio, CI = Confidence Interval, sdHR=Sub-distribution Hazard Ratio, EPTB = extra pulmonary tuberculosis, PTB = pulmonary tuberculosis, Both = extra pulmonary tuberculosis and pulmonary tuberculosis, RmR-TB = Rifampicin monoresistant tuberculosis, Pre-XDR-TB = Pre-extensively drug resistant tuberculosis

**Table 4:**
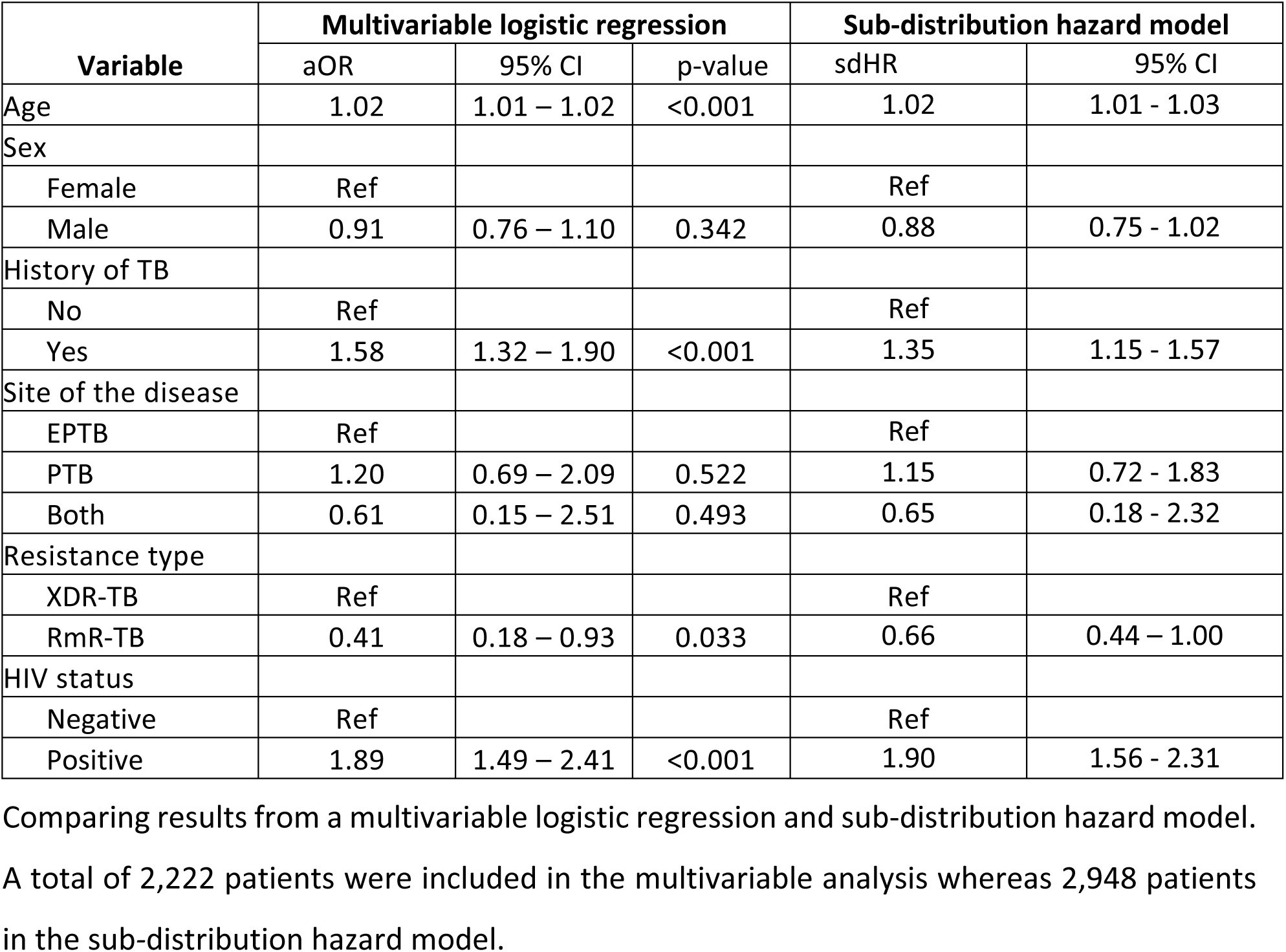
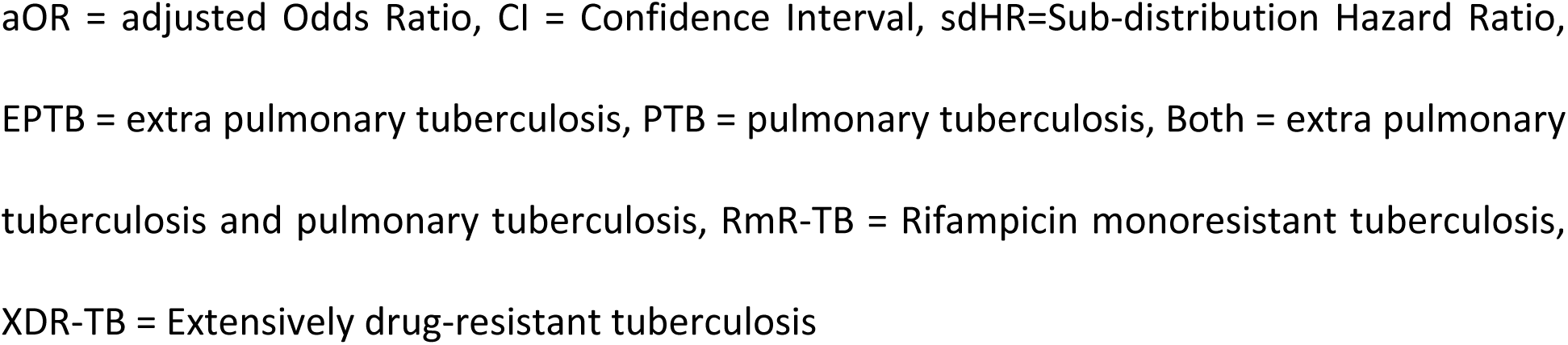
Two-year mortality among patients with rifampicin mono-resistant and extensively drug-resistant tuberculosis.

| Variable | Multivariable logistic regression |  |  | Sub-distribution hazard model |  |
| --- | --- | --- | --- | --- | --- |
|  | aOR | 95% CI | p-value | sdHR | 95% CI |
| Age | 1.02 | 1.01 – 1.02 | <0.001 | 1.02 | 1.01 - 1.03 |
| Sex |  |  |  |  |  |
| Female | Ref |  |  | Ref |  |
| Male | 0.91 | 0.76 – 1.10 | 0.342 | 0.88 | 0.75 - 1.02 |
| History of TB |  |  |  |  |  |
| No | Ref |  |  | Ref |  |
| Yes | 1.58 | 1.32 – 1.90 | <0.001 | 1.35 | 1.15 - 1.57 |
| Site of the disease |  |  |  |  |  |
| EPTB | Ref |  |  | Ref |  |
| PTB | 1.20 | 0.69 – 2.09 | 0.522 | 1.15 | 0.72 - 1.83 |
| Both | 0.61 | 0.15 – 2.51 | 0.493 | 0.65 | 0.18 - 2.32 |
| Resistance type |  |  |  |  |  |
| XDR-TB | Ref |  |  | Ref |  |
| RmR-TB | 0.41 | 0.18 – 0.93 | 0.033 | 0.66 | 0.44 – 1.00 |
| HIV status |  |  |  |  |  |
| Negative | Ref |  |  | Ref |  |
| Positive | 1.89 | 1.49 – 2.41 | <0.001 | 1.90 | 1.56 - 2.31 |

Comparing results from a multivariable logistic regression and sub-distribution hazard model. A total of 2,222 patients were included in the multivariable analysis whereas 2,948 patients in the sub-distribution hazard model.
aOR = adjusted Odds Ratio, CI = Confidence Interval, sdHR=Sub-distribution Hazard Ratio, EPTB = extra pulmonary tuberculosis, PTB = pulmonary tuberculosis, Both = extra pulmonary tuberculosis and pulmonary tuberculosis, RmR-TB = Rifampicin monoresistant tuberculosis, XDR-TB = Extensively drug-resistant tuberculosis

The cumulative incidence of mortality in the first four months (120 days) of treatment was highest for patients with RmR-TB compared to other resistance types (MDR-TB, pre-XDR-TB, and XDR-TB). Thereafter, the highest mortality was observed in people with XDR-TB (Fig 4).

**Fig 4:**
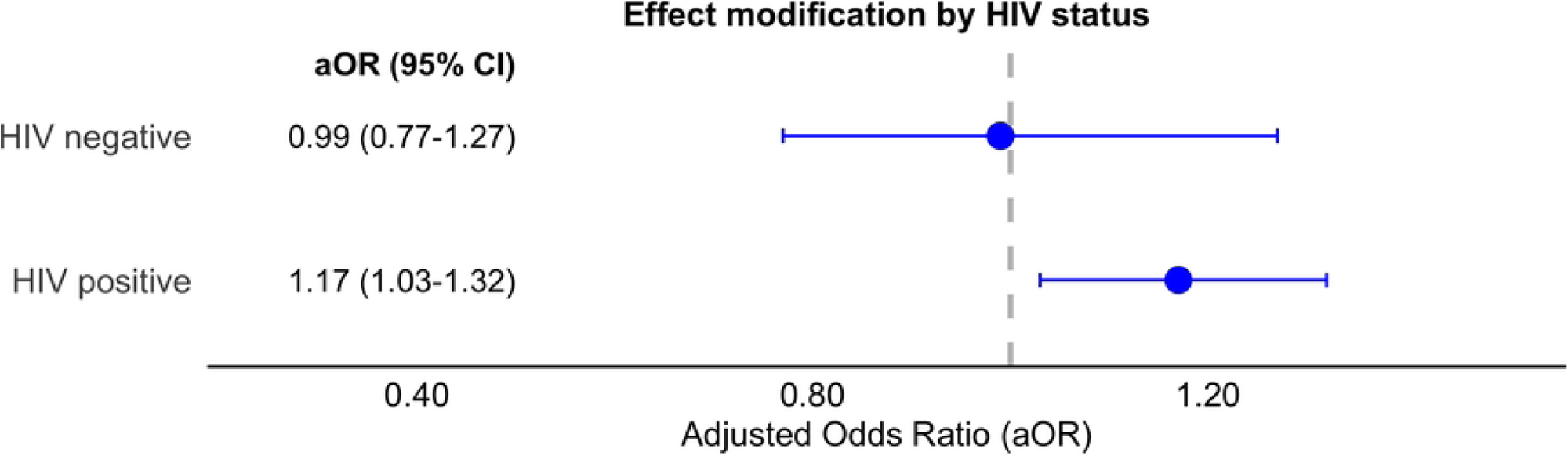
Cumulative incidence of two-year mortality stratified by resistance type. **RmR-TB** = Rifampicin monoresistant tuberculosis, MDR-TB = multi-drug-resistant tuberculosis, Pre-XDR-TB = pre-extensively drug-resistant tuberculosis, XDR-TB = Extensively drug-resistant tuberculosis

## Discussion

The increasing burden of rifampicin mono-resistant tuberculosis (RmR-TB) is a growing concern in many regions of the world. The higher risk of mortality posed by RmR-TB compared to MDR-TB has been hypothesized to be due to HIV co-infection, but prior studies were not powered to assess this hypothesis. In this large global IPD meta-analysis, we found that the odds of mortality are indeed higher among patients with RmR-TB than those with MDR-TB and that the effect does not vary by HIV status even though the odds of mortality remained higher among those with HIV co-infection.

Previous studies primarily focused on the epidemiology and risk factors associated with RmR-TB, with few studies addressing treatment outcomes [2, 3, 5]. A retrospective cohort study in California reported a two-fold increase in mortality risk among RmR-TB patients compared to those with drug-susceptible TB [27]. Conversely, a study in South Africa found lower odds of mortality between patients with RmR-TB or MDR-TB compared to those with fluoroquinolone resistance [28]. However, the latter included patients diagnosed with RR-TB by Xpert MTB-RIF without further drug susceptibility testing. These two studies had small sample sizes, limiting their power to compare mortality risk between RmR-TB and other types of DR-TB [16]. The robustness of our results of increased risk of mortality among patients with RmR-TB is demonstrated by a statistically significant effect even when accounting for loss to follow-up as a competing risk.

Previous studies have suggested that HIV may contribute to the higher risk of mortality among patients diagnosed with RmR-TB. The stratified analysis showed a higher risk of mortality of RmR-TB among those with HIV co-infection but not among HIV-negative patients. Nevertheless, HIV co-infection did not statistically modify the relationship between RmR-TB and mortality, indicating that factors beyond HIV status are likely influencing the increased mortality risk in RmR-TB patients. Existing evidence highlights other contributing factors, including history of TB treatment, comorbidities, nutritional status, and treatment adherence, all of which can elevate mortality risk, particularly in resource-limited settings with a higher burden of HIV [3, 14, 16]. Our findings revealed that mortality was higher among RmR-TB patients previously treated or TB supporting findings from previous studies.

The strength of this study lies in the fact that we used a large patient dataset covering four WHO regions, allowing for a robust comparison not only between RmR-TB and MDR-TB but also against pre-XDR-TB and XDR-TB. Additionally, our sensitivity analysis including LTFU as a competing risk, enhancing the reliability of our findings. Despite these strengths, our study findings should be interpreted considering the following limitations. First, we only adjusted for baseline DST results, while patients may acquire additional resistance throughout the treatment course. Second, the variability of the diagnostic tools used to derive DST results may have led to differential exposure misclassification as the sensitivity and specificity differ by the diagnostic tool used [10, 12]. As resistant TB strains circulate and mutate at different rates across regions, the observed estimates of mortality may be skewed[29]. Furthermore, implementing a complete case analysis may introduce bias due to missing data on important covariates such as BMI, anti-retroviral treatment use, time between TB diagnosis and treatment initiation, prior treatment regimen, and adherence. The potential for residual confounding could also raises concerns about the robustness of our conclusions.

In conclusion, our findings highlight the high risk of mortality among patients diagnosed with RmR-TB, especially within the first four months of treatment. The increased odds of mortality among PLHIV underscores the need for timely HIV diagnosis and early initiation of ART as this can reduce the overall mortality of TB. Further research is needed to elucidate the biological and social determinants contributing to the higher burden of RmR-TB among those with HIV co-infection and the reason for high mortality.

Key areas for investigation include the role of immune suppression, drug resistance patterns, and the impact of comorbidities on treatment outcomes. Furthermore, strengthening surveillance systems to monitor RmR-TB trends and outcomes among those with HIV co-infection, is essential to inform targeted interventions and resource allocation. Addressing these gaps will be essential for improving survival rates and optimizing the management of drug-resistant TB in this vulnerable population.

## Data Availability

We have no authority to share the data however, this data can be accessed from the TBIPD platform and the TB Consortium portal. Individual datasets used can be accessed from individual authors upon request.

https://www.ucl.ac.uk/global-health/research/z-research/tb-ipd-platform

https://tbportals.niaid.nih.gov

## Acknowledgements

Data were obtained from the TB Portals (https://tbportals.niaid.nih.gov), which is an open-access TB data resource supported by the National Institute of Allergy and Infectious Diseases (NIAID) Office of Cyber Infrastructure and Computational Biology (OCICB) in Bethesda, MD. These data were collected and submitted by members of the TB Portals Consortium (https://tbportals.niaid.nih.gov/Partners). Investigators and other data contributors who originally submitted the data to the TB Portals did not participate in the design or analysis of this study. We would also like to acknowledge Kathryn Schnippel for additional data provision.

